# Evaluating the suitability of the case-crossover design under changing baseline outcome risk: A simulation of ambient temperature and preterm birth

**DOI:** 10.1101/2021.02.17.21251948

**Authors:** Daniel Carrión, Johnathan Rush, Elena Colicino, Allan C. Just

## Abstract

Case-crossover (CCO) studies are case-only within-person analyses of associations between acute exposures and outcomes. By design, the CCO is not confounded by time-invariant characteristics. CCO studies assume stable baseline outcome risks. Since the baseline risk of birth increases secularly over gestation, the validity of CCOs of preterm birth (PTB) is not clear. We simulated associations between temperature and PTB in New York State using historical ambient temperature data for LaGuardia Airport and PTB data from the CDC using different control period durations. CCO analyses were conducted with conditional logistic regression. We calculated bias according to the absolute difference between the simulated and estimated effects in the natural log scale, and found that 2-week, 28-day, and 1-month stratified control period selection yielded negligible bias across all simulated effects. Coverage of the 95% confidence intervals was also appropriate across all three control selection strategies. Our findings suggest that the time-stratified CCO should yield appropriate inference in studies of ambient temperature and preterm birth.

## Background

The case-crossover (CCO) design is widely used in epidemiology. CCO studies are case-only, within-person comparisons, therefore not confounded by time-invariant characteristics.^1^ Proper statistical inference with CCO studies relies on appropriate selection of control time periods, and the time-stratified design is a preferred method for control selection.^2^ However, CCO studies assume stable baseline outcome risks.^3^ Multiple studies have used the CCO approach linking ambient temperature and preterm birth (PTB),^4^ but since the baseline PTB risk increases secularly over gestation, the validity of the CCO design for PTB is unclear.^5^ We utilized simulation methods to assess the appropriateness of the CCO in associations between ambient daily temperature and PTB.

## Methods

We conducted a simulation using 2018 data for New York State. Data were acquired from National Weather Service records for LaGuardia Airport (daily maximum temperature) and the Centers for Disease Control’s WONDER database (PTB data).^6^ Daily births by gestational age are unavailable so were estimated using marginal distributions of births by month and day of week (within month).

Estimated daily births per gestational age (20-36 weeks) served as the basis for baseline risk. Baseline risks were combined with simulated (true) temperature effects to create expected counts per day. Relative risks (RR) were set from 0.9 to 1.25 per 10°F increase on lag day 0. Repeated Poisson random number generation created 1000 datasets per simulated RR.^7^ Counts were disaggregated into individual records for CCO analyses, estimated via conditional logistic regression with 2-week, 28-day, and 1-month stratified control period selection matched on the day of week. Analyses only included warm months, May through September.^4^ A sensitivity analysis included modeling 2007 data.

We summarized results according to bias and coverage. We calculated bias according to the absolute difference 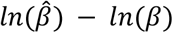, where 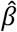 is the estimated effect and*β*is the true effect.Coverage is the proportion of simulations where the 95% confidence intervals include the true effect. Reproducible analyses were conducted in R 4.1.0 and using the *targets* package.^8^

## Results

All three control-selection strategies exhibited little bias (**Figure 1A**). There was a median bias, across all simulated effects, of 0 for both the 2-week and 28-day stratified results. The month results exhibited a median bias of −0.001. The 2-week results exhibited the largest variability as measured by the interquartile range (0.023), compared to the other two models, which both had interquartile ranges of 0.016.

**Figure 1:**
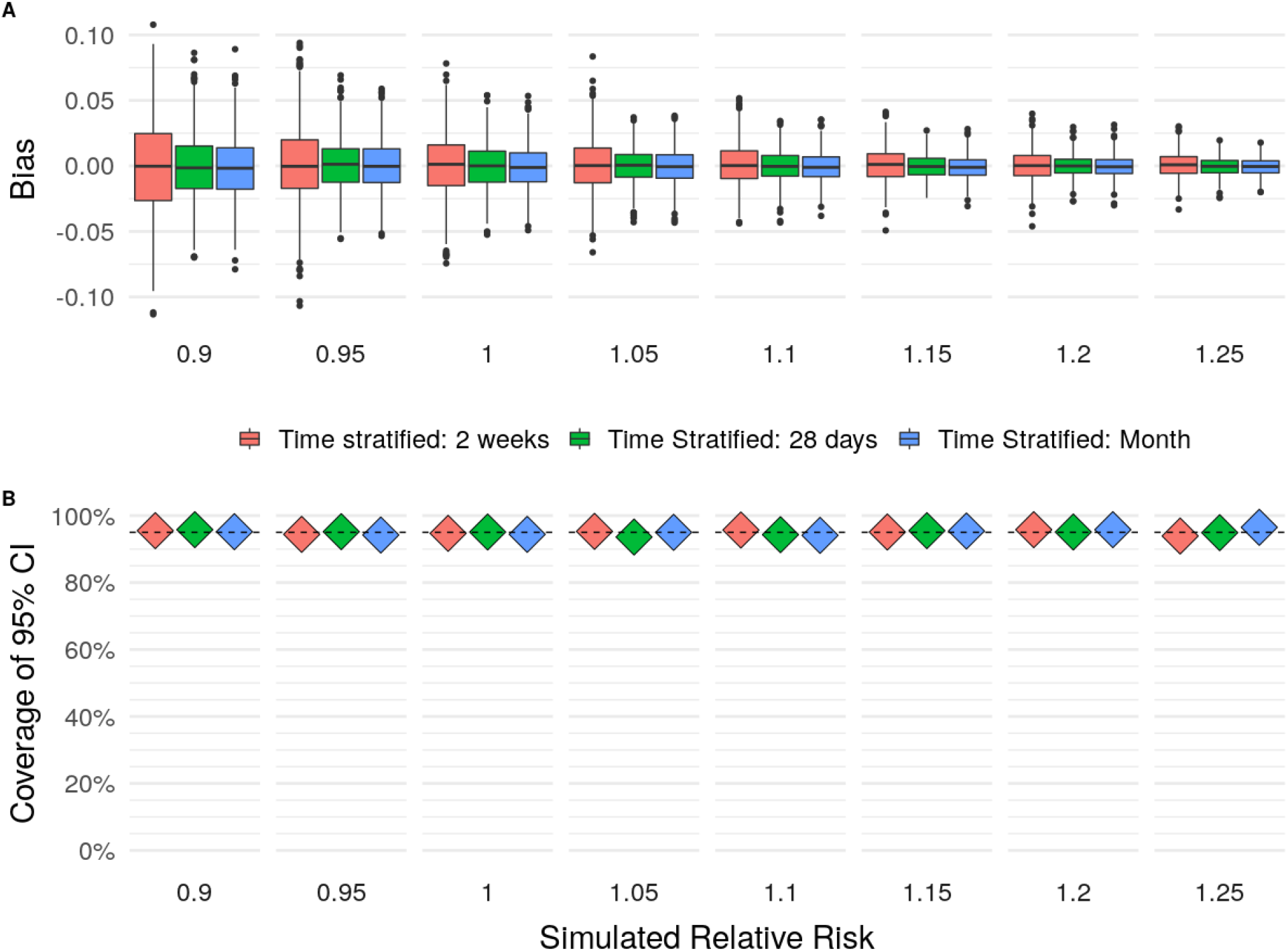
Results from 2018 simulations and case-crossover analyses. Colors represent analysis type, namely 2-week stratified, 28-day stratified, and 1-month stratified control selection. **A**) Distribution of bias for each simulated effect, scaled to a 10F increase in temperature. **B**) The proportion of 1,000 simulations where confidence intervals contain the simulated effect. Dashed line at 95%.

Coverage of 95% confidence intervals was consistent for all control selection strategies; between 94-96%, 94-97%, and 94-96% of all intervals included the simulated effect for 2-week, 28-day, and month-stratified results respectively (**Figure 1B**). Sensitivity analyses simulating data from 2007 were consistent in bias and coverage (Supplemental materials).

## Conclusions

CCO studies may suffer systematic bias if selected control times are non-exchangeable within person.^1^ While temperature varies seasonally and the risk of PTB changes quickly over pregnancy, our simulations show that the CCO study design yields negligible bias and appropriate coverage of confidence intervals. This is important because many studies in perinatal epidemiology have used 28-day and month stratification approaches, but no studies have yet assessed the appropriateness of the CCO in this context. We also tested a 2-week approach as an alternate strategy to evaluate the consistency of results under concerns of rapidly changing baseline risks, thus using narrower time strata. These results were consistent, but yielded a wider range of estimates. Overall, our study supports the applicability of the CCO for ambient environmental exposures and preterm birth.

## Data Availability

All data and analytical code to reproduce results are available on GitHub.

https://github.com/justlab/casecrossover_preterm_simulation

## Data availability

All data and analytical code to reproduce results are available on GitHub: https://github.com/justlab/casecrossover_preterm_simulation.

