## Supplemental Materials for "Evaluating the suitability of the case-crossover design under changing baseline outcome risk: A simulation of ambient temperature and preterm birth"

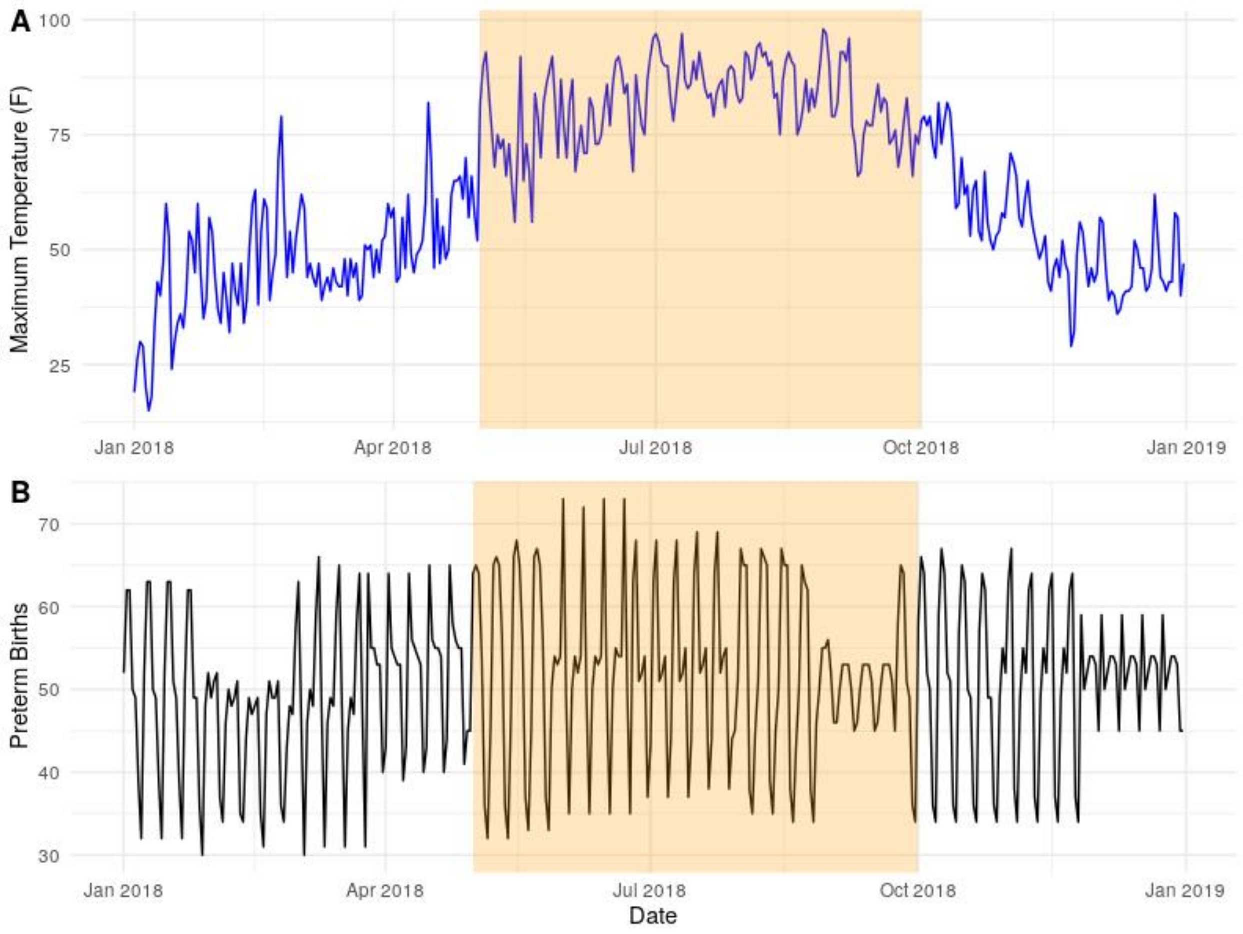


**Supplemental Figure 1: Data inputs for simulations and subsequent case crossover analyses, 2018.** A) Maximum daily temperature from LaGuardia Airport records. B) Estimated daily preterm births across all gestational ages 20-36 weeks in New York State. The highlighted section represents warmer months modeled in case-crossover analysis.

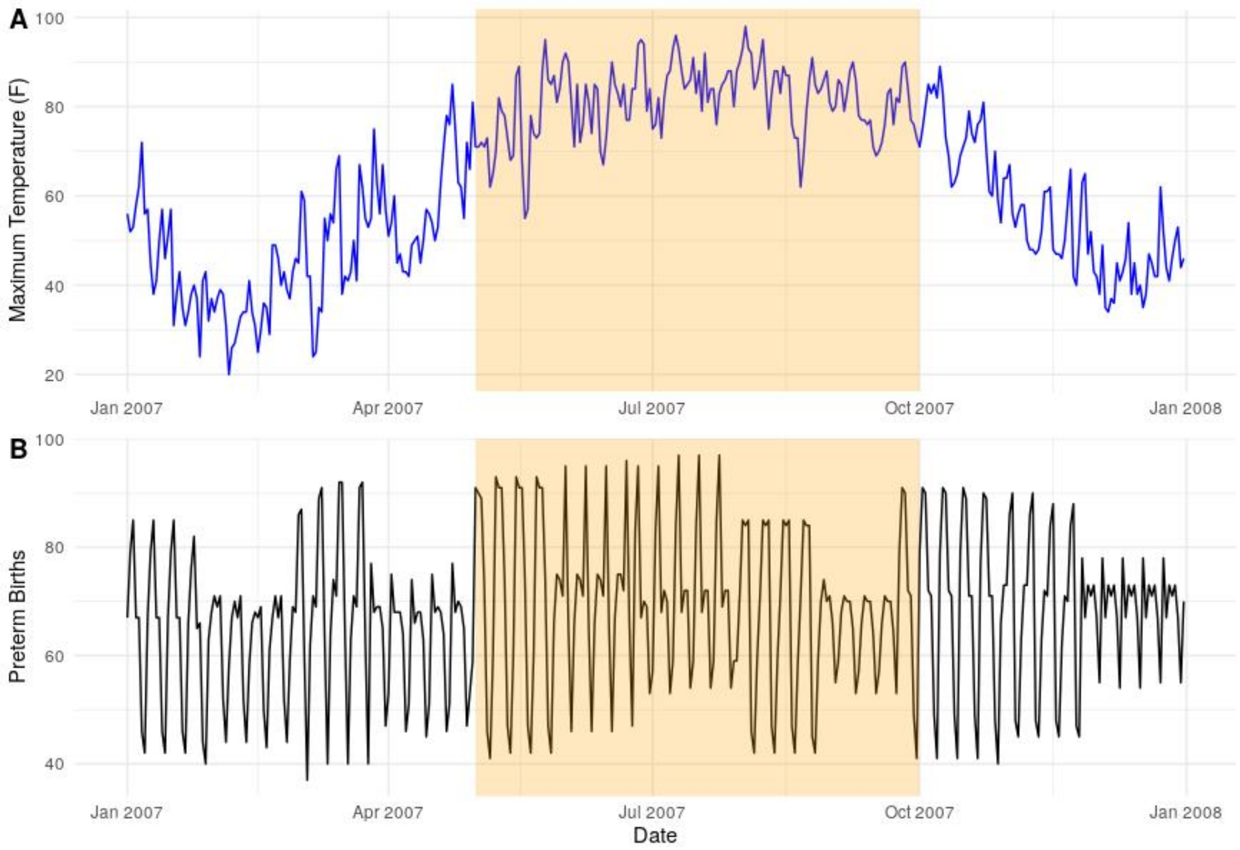


**Supplemental Figure 2: Data inputs for simulations and subsequent case crossover analyses, 2007.** A) Maximum daily temperature from LaGuardia Airport records. B) Estimated daily preterm births across all gestational ages 17-36 weeks in New York State. The highlighted section represents warmer months modeled in case-crossover analysis.

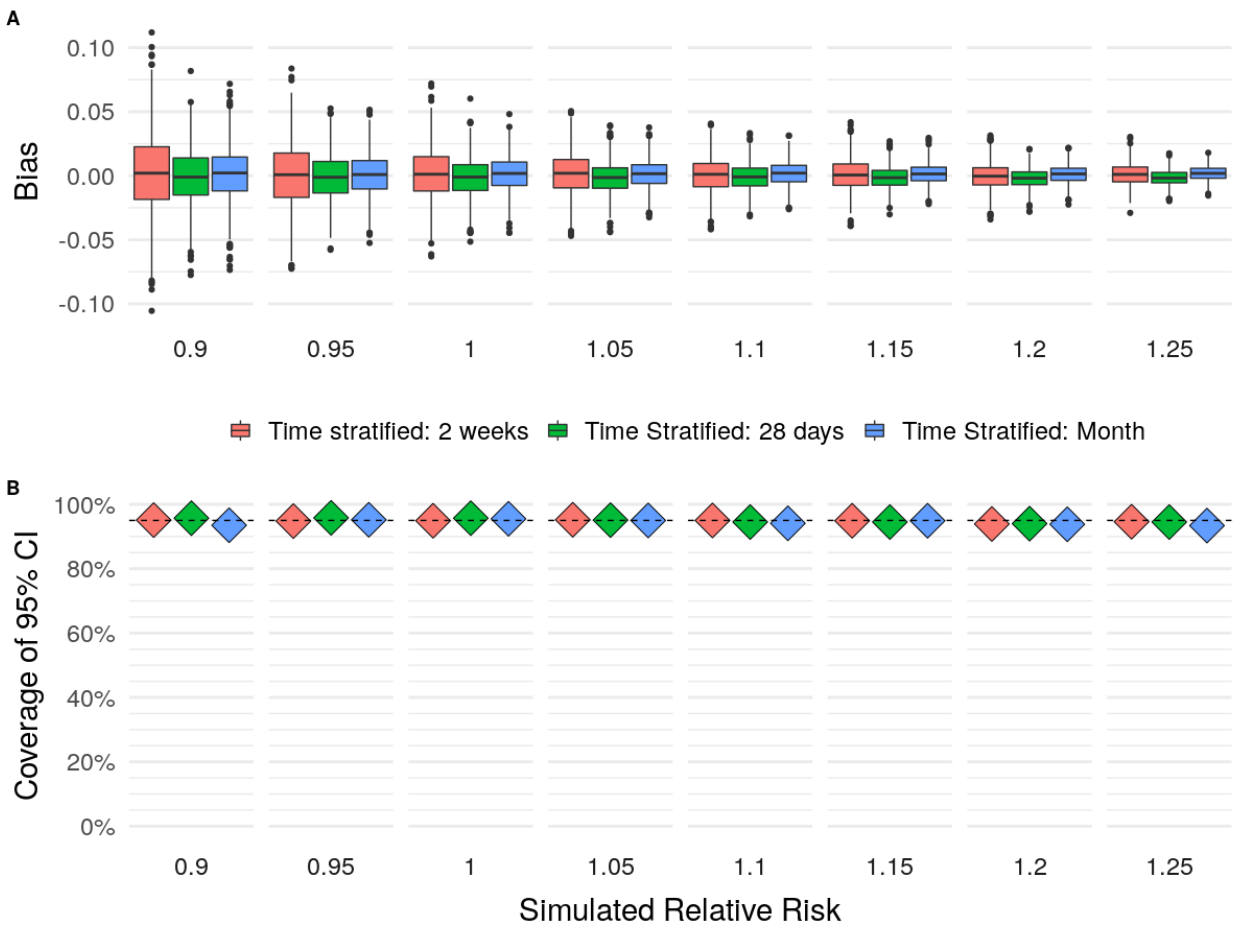


**Supplemental Figure 3: Results from 2007 simulations and case-crossover analyses.** Colors represent analysis type, namely 2-week stratified, 28-day stratified, and 1-month stratified control selection. **A**) Distribution of bias for each simulated effect, scaled to a 10F increase in temperature. **B**) The proportion of 1,000 simulations where confidence intervals contain the simulated effect. Dashed line at 95%.
